# Incremental Value of Cerebrospinal Fluid Biomarker-Integrated Classification of Cerebral Amyloid Angiopathy

**DOI:** 10.64898/2026.08.30.26361511

**Authors:** Mattia Losa, Matteo Cotta Ramusino, Ilaria Gandoglia, Federico Mazzacane, Beatrice Orso, Luigi Lorenzini, Andrea Donniaquio, Federico Massa, Elena Sentieri, Lorenzo Gualco, Giulia Perini, Valentino De Franco, Alfredo Costa, Francesco Bax, Steven M. Greenberg, Mariel G. Kozberg, Fabrizio Piazza, Antonio Uccelli, Angelo Schenone, Massimo Del Sette, Lisa Maria Farina, Luca Roccatagliata, Matteo Pardini

## Abstract

**Background:** The Boston Criteria v2.0 represent the gold standard for diagnosing Cerebral Amyloid Angiopathy (CAA), but their application is currently precluded in mixed small vessel disease (SVD), where deep and lobar hemorrhages coexist. The aims of this study are: (i) to determine which cerebrospinal fluid (CSF) biomarker (Aβ42, Aβ40, Aβ42/40 ratio) is the best candidate to support the CAA diagnosis; (ii) to define a data-driven cut-off, and (iii) to explore if a biomarker-integrated classification significantly improves the phenotypical concordance with the suspected predominant SVD (CAA vs. arteriosclerosis).

**Methods:** We analyzed data from a retrospective multicenter cohort of patients with suspected CAA, defined as probable CAA (Boston criteria v2.0) but allowing deep hemorrhagic lesions, and with available CSF biomarkers. We visually quantified MRI-visible SVD markers (e.g., cerebral microbleeds [CMB], cortical superficial siderosis [cSS], lacunes) and their association with MRI-visible SVD features. We employed a Gaussian Mixture Model (GMM) to identify a data-driven threshold for amyloid positivity (A^+^). Then, we compared the prevalence of MRI-visible manifestations of SVD between subgroups applying different frameworks, namely the current MRI-based classification (probable CAA vs. mixed SVD) and a CSF biomarker-integrated classification (A^+^ vs. A^-^).

**Results:** We enrolled 121 patients (age: 72 [66-77] years; 60% probable CAA, 40% mixed SVD with suspected CAA). The CSF Aβ42/40 ratio showed a bimodal distribution and consistent associations with all CAA-specific radiological features. The CSF biomarker-integrated reclassification, particularly using the GMM cut-off, significantly improved the distinction between subgroups regarding CAA- and arteriosclerosis-related MRI features (e.g., cSS presence: probable CAA vs. mixed SVD: aOR=2.84 [95%CI 1.27-6.39], p=0.011; A+ vs. A-: aOR=12.68 [95%CI 4.31-37.32], p<0.001; deep lacunes presence: probable CAA vs. mixed SVD: aOR=0.20 [95%CI 0.08-0.50], p<0.001; A^+^ vs. A^-^: aOR=0.04 [95%CI 0.01-0.11], p<0.001). Notably, patients classified as A^+^ never demonstrated more than four deep CMBs.

**Discussion:** A CSF biomarker-integrated classification may improve the classification of CAA compared with the current MRI-based framework. These findings are cohort-specific and would benefit from further validation, especially with a neuropathological reference. Still, these results support a future transition toward an integrated biological-radiological framework, which may refine in vivo CAA diagnosis, particularly in mixed SVD.

## 1. Introduction

Cerebral amyloid angiopathy (CAA) is one of the most common cerebral small vessel diseases ( SVD)^1^. Cortical and leptomeningeal vascular beta-amyloid (Aβ) deposition can be suspected in-vivo in the presence of a characteristic radiological pattern^2^. Accordingly, the MRI-visible manifestations of the disease have been incorporated into diagnostic criteria, which have progressively advanced over the years the ability to detect CAA in vivo^3^. However, even in the latest iteration (Boston Criteria v2.0), diagnostic accuracy remains imperfect, mainly because no validated disease-specific biomarker is currently available, unlike in other related disorders where biological evidence is now required for an accurate diagnosis^4–6^

The Boston criteria v2.0 are mainly based on strictly lobar hemorrhagic MRI markers in an appropriate clinical context, with deep hemorrhagic lesions serving as exclusion criteria^3^. Patients with both lobar and deep hemorrhagic markers are therefore usually classified as having mixed-SVD, a pattern often associated with arteriosclerosis or deep perforator arteriopathy (DPA). Nevertheless, mixed-SVD is heterogeneous, and underlying CAA may exist in a subset of these patients^7–9^. In clinical practice, CAA may still be suspected when mixed-SVD is accompanied by supportive imaging features, particularly a predominantly lobar hemorrhagic burden and/or cortical superficial siderosis (cSS)^10^. Conversely, some patients classified as probable CAA may be false-positive cases, reflecting predominant arteriolosclerosis rather than amyloid angiopathy^11^. In this context, biological support for the diagnosis would be advantageous to rule in (or rule out) the diagnosis (**Figure 1**). Even if evidence about a CAA-like Cerebrospinal Fluid (CSF) biomarker profile is growing at the group level, this molecular signature is not clearly standardized, lacks a CAA-specific cut-off, and has not yet been demonstrated to improve diagnostic accuracy^12^. Relying on an integrated radiological and biological framework has the potential to improve the detection of the predominant underlying type of SVD responsible for the clinical picture (i.e., CAA versus DPA), with subsequent clinical impact for prognosis prediction and antithrombotic therapies.

**Figure 1.**
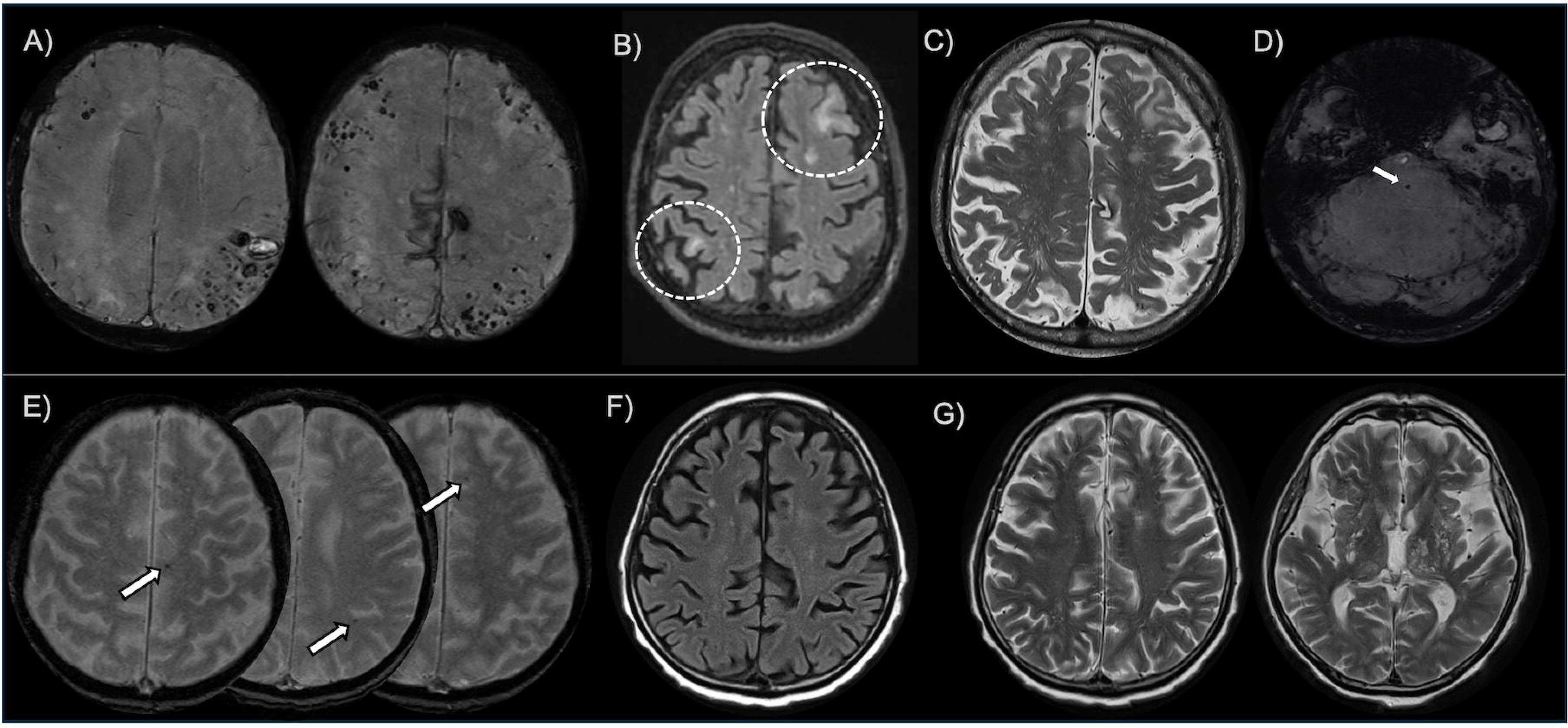
Clinically challenging scenarios of patients with suspected CAA. In the upper row, a male patient in his 70s, with multiple vascular risk factors, admitted for subacute cognitive decline with a radiological picture highly suspicious for CAA, who does not fulfil Boston criteria given the presence of a single deep (pontine) microbleed. CSF biomarkers showed an A+T+N+ profile, according to the ATN framework. In the lower row, a male patient in his 60s, with hypertension and an amnestic MCI, fulfilling the “probable CAA” definition but without evidence of Aβ deposition (ATN classification: A-T-N-). A) SWI sequences with evidence of multiple lobar CMBs, with a greater cluster in the left parieto-occipital lobe, a small acute ICH, and cortical superficial siderosis in the cingulate cortex bilaterally. B) Multifocal hyperintensities in FLAIR sequences locally associated with multiple CMBs suggestive of CAA-related inflammation. C) Severely enlarged perivascular spaces of the centrum semiovale. D) Single pontine CMB. E) Multiple slices of GRE sequence with strictly lobar CMBs (total count: 5). F) FLAIR sequence with low burden of WMH (Fazekas scale: 1); G) T2-weighted sequence of the patient with moderate enlargement of the CSO-PVS, but with predominant enlargement of the BG-PVS. Legend: A=amyloid CAA=cerebral amyloid angiopathy; CMB=Cerebral Microbleeds; CSF=Cerebrospinal Fluid; cSS=cortical superficial siderosis; ICH=Intracerebral Hemorrhage; MCI=Mild Cognitive Impairment N=Neurodegeneration; T=Tauopathy

This study aims to evaluate whether integrating CSF biomarkers into the CAA framework provides a more accurate classification than the current MRI-based approach. The main aims of the study are: (i) to determine which CSF biomarkers (Aβ42, Aβ40, or Aβ42/40 ratio) is the optimal candidate as a supportive biomarker for the CAA diagnosis; (ii) to establish a data-driven cut-off, moving beyond thresholds designed for Alzheimer’s disease (AD); (iii) to explore if an integrated biological and radiological framework applied in patients with suspected CAA significantly improves phenotypical concordance with the suspected predominant underlying SVD.

## 2. Materials and Methods

### 2.1 Ethical Approval and Patient Consent

All procedures contributing to this work comply with the ethical standards of the relevant national and institutional committees on human experimentation and with the Helsinki Declaration of 1975, as revised in 2008. Written informed consent was obtained from all participants as part of an observational protocol approved by the Ethics Committee. The manuscript was prepared with reference to the Strengthening the Reporting of Observational Studies in Epidemiology (STROBE) guidelines.

### 2.2 Study participants and inclusion/exclusion criteria

This is a multicentric observational retrospective study conducted between January 2019 and December 2025 at two Italian research hospitals: the IRCCS Azienda Ospedaliera Metropolitana (AOM), San Martino Hospital (Genoa), and the IRCCS C. Mondino Foundation (Pavia).

All patients fulfilled the following inclusion criteria: (i) patients fulfilling Boston criteria v2.0 for probable CAA, or (ii) patients with an otherwise probable CAA phenotype but concomitant deep hemorrhagic lesions, who would not fulfill Boston v2.0 criteria because of these lesions; (iii) patients accepted to perform during diagnostic work-up a lumbar puncture for CSF biomarkers analysis within 6 months from diagnosis (baseline MRI). For simplicity, the included patients will be defined from now on as “suspected CAA”, to avoid confusion with the “possible” or “probable CAA” category of the Boston criteria framework. Exclusion criteria were: (i) motion artifacts or absence of sequences required to assess Boston criteria; (ii) contraindications or refusal to perform lumbar puncture; (iii) suspected or confirmed CAA mimics (e.g., radiotherapy, brain tumors, critical illness-associated microbleeds, ischemic stroke with hemorrhagic transformation); (iv) monogenic SVD. CSF analysis was offered to all suspected CAA patients during this period as part of the initial diagnostic work-up, based on clinical indication (e.g., cognitive decline, exclusion of differential diagnoses, or to support diagnosis). Of note, neither iatrogenic CAA nor CAA-related inflammation (CAA-ri) was considered an exclusion criterion^13,14^. These inclusion and exclusion criteria were selected to approximate a real-world clinical setting in which MRI-visible SVD markers may reflect CAA, DPA, or their coexistence.

We collected the demographics, medical, and medication history of enrolled subjects. Clinical presentations were divided into the following categories: hemorrhagic presentation (ICH or cSAH), transient focal neurological episodes (TFNE), or cognitive presentation (mild cognitive impairment, MCI, or dementia). Global cognitive performance was evaluated using the Mini-Mental Status Examination (MMSE).

### 2.3 MRI acquisition and analysis

Baseline MRI scans (1.5 or 3 Tesla) were first evaluated for diagnostic purposes by two neurologists with expertise in neuroimaging and MRI ratings (ML and MCR), who verified the inclusion/exclusion criteria. Patients were classified as probable CAA or mixed SVD, according to Boston criteria v2.0^15–17^.

Subsequently, the rating of the MRI images was performed locally by ML (in Genoa) and MCR (in Pavia), blinded to CSF biomarker profiles and clinical data. In cases of uncertainty during the rating process, images were re-examined with a senior neuroradiologist at the respective site (LR in Genoa and LMF in Pavia) to achieve a final consensus. Hemorrhagic and non-hemorrhagic MRI-visible manifestations of cSVD were evaluated according to updated STRIVE^18^. Blood-sensitive sequences used to evaluate hemorrhagic features were T2* gradient echo (T2* GRE) sequences or susceptibility-weighted imaging (SWI). Hemorrhagic lesions were visually classified as follows: number of CMBs (according to MARS classification, hemorrhagic lesions < 10mm diameter)^19^, were visually counted and assessed as low (<3 CMBs), mild (between 4 and 10 CMBs), or high burden (>10 CMBs) for lobar CMB, while low (1 CMB), moderate (2-3 CMB) and high burden (>3) for deep CMBs. cSS was classified as present or absent and then scored according to the cSS multifocality score^20^. White matter hyperintensities (WMH) were scored according to the Fazekas scale on T2-FLAIR sequences^21^. Centrum semiovale and basal ganglia enlarged perivascular spaces (CSO- and BG-EPVS) were detected on T2-weighted sequences, rated as previously proposed^22^. The presence of deep lacunes was rated on T2-FLAIR sequences^18^. Lobar intracerebral hemorrhages (ICH: hemorrhagic lesions ≥ 10 mm) were identified and assessed as present or absent. Then, two composite small vessel disease score were assessed: the CAA-SVD score (range from 0-6), as previously described^23^, and the DPA-SVD score (range 0-4), composed by the Fazekas scale (1 point if deep WMH>1), BG-PVS (1 point if >20), deep lacunes (1 point if present) and deep CMB (1 point if present)^24^.

### 2.4 CSF collection and analysis

Following standard operating procedures, CSF samples (6 to 8 mL) were obtained via lumbar puncture at the L3-L4 or L4-L5 interspace in the early morning^25^. The CSF was collected using sterile polypropylene tubes, then centrifuged at 4000 g for 10 minutes at 4°C. To ensure the long-term stability of proteins, the resulting aliquots were stored in polypropylene tubes at −80°C until analysis. We utilized the same assay in the two centers, the Lumipulse G600 II® fully automated chemiluminescent enzyme immunoassay system (Fujirebio Europe, Gent, Belgium) to measure core CSF biomarkers (Aβ42, Aβ40, p-Tau181, and t-Tau). Intra-test reproducibility had an average coefficient of variation (CV) below 10%, and inter-test reproducibility averaged below 15% for all assays. Assay cartridge datasheet cutoffs were also verified in our centers: 600 pg/mL for Aβ42; 0.069 pg/mL for Aβ42/40 ratio; 404 pg/mL for total-Tau; 56.5 pg/mL for pTau^26^. Intratest reproducibility had an average coefficient of variation of <10%, and intertest reproducibility averaged <15% for all assays.

### 2.5 Statistical analysis

Descriptive statistics about clinical, radiological, and CSF biomarker data were expressed as mean, standard deviation (SD), or median and interquartile range (IQR), as appropriate. Normality was assessed using the Shapiro-Wilk test and histograms. We compared subgroups using Student’s or Mann-Whitney tests for continuous variables and Chi-squared or Fisher’s exact tests for categorical variables, as appropriate.

We first aimed at exploring CSF biomarker (Aβ42, Aβ40, and Aβ42/40 ratio) distribution and their association with MRI-visible manifestations of SVD. Specifically, we applied multivariable binomial and ordinal logistic regression models (adjusted for age, sex, centers, and MRI protocol); to compare the effect size of the association, we normalized the CSF biomarker values using z-score normalization. Biomarkers with a binomial distribution and more consistent associations with the CAA radiological signature were considered the reference for the subsequent analysis.

We then aimed at defining a data-driven threshold to overcome the possible limitations of AD-related cut-offs in this clinical scenario. To do so, we applied an unsupervised Gaussian Mixture Modeling (GMM) on the CSF biomarker to detect an optimal distinction point between “amyloid-positive” (A^+^) and “amyloid-negative” (A^-^) biological populations. This approach yielded an MRI-independent patient classification strictly driven by the cohort’s inherent biochemical properties.

Third, to evaluate the possible impact of incorporating CSF biomarkers into SVD classifications, we divided patients with suspected CAA following three frameworks: (i) the current MRI-based classification (Boston criteria v2.0 – probable CAA vs. mixed SVD)^3,27^; then, we reclassified patients into MRI-independent subgroups based on the evidence of Aβ deposition using (ii) the AD-standard cut-off and (iii) the data-driven cut-off. The three different classifications were explored using multivariable logistic regression models (dependent variable: radiological marker; explanatory variable: classification status), adjusted for age, center, and MRI protocol. We hypothesized that an increase in phenotypical radiological differences between groups would represent an indirect indicator of meaningful reallocation of patients between frameworks. The p-for-interaction was used to verify whether biomarker-integrated models yielded significantly superior associations with SVD features compared to the standard diagnostic criteria. Finally, we performed pre-specified sensitivity analyses, excluding i) patients with evidence of AD-related tauopathy and ii) patients with evidence of CAA-ri, a possible/probable iatrogenic CAA, and/or patients who performed the lumbar puncture less than a month from a symptomatic ICH.

Statistical analysis was performed using Jamovi (version 2.6.17; https://www.jamovi.org) and R (version 2025.05.1; http://www.r-project.org/ - “mclust” package for GMM)^28,29^. The threshold for statistical significance was set to p<0.05.

## 3. Results

### 3.1 Study population

One-hundred twenty-one patients with suspected CAA were enrolled in the study, of which 74 patients had a probable CAA, and 47 had mixed SVD (**Table 1**). A flowchart of patient selection is provided in **eFigure1**. The majority of the cohort exhibited a cognitive onset (n=62, 51%). To note, a subset of patients (n=15, 12%) exhibited an index clinical presentation not included in the Boston criteria (e.g., ischemic stroke, parkinsonism), but with concomitant cognitive impairment, thereby fulfilling the Boston criteria. About CSF biomarkers, probable CAA showed lower Aβ42 (371 [315-550] vs. 542 [356-887]; p=0.006), Aβ42/40 ratio (0.05 [0.04-0.06] vs. 0.09 [0.05-0.10]; p<0.001), and higher pTau181 levels (60 [38-80] vs. 32 [22-58]; p<0.001) compared to mixed SVD, while no difference was observed in Aβ40 and total-Tau levels (p>0.05).

**Table 1.**
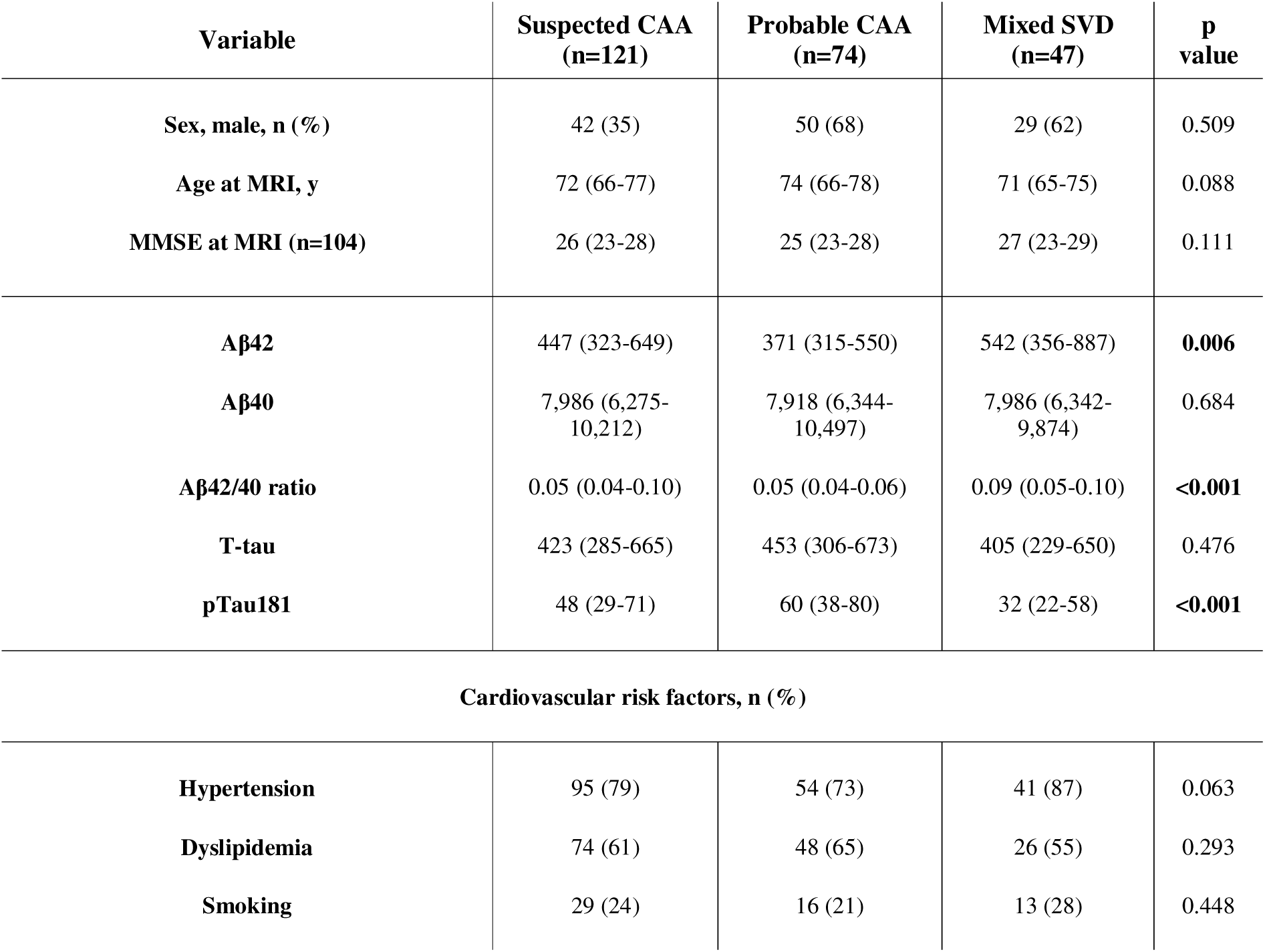
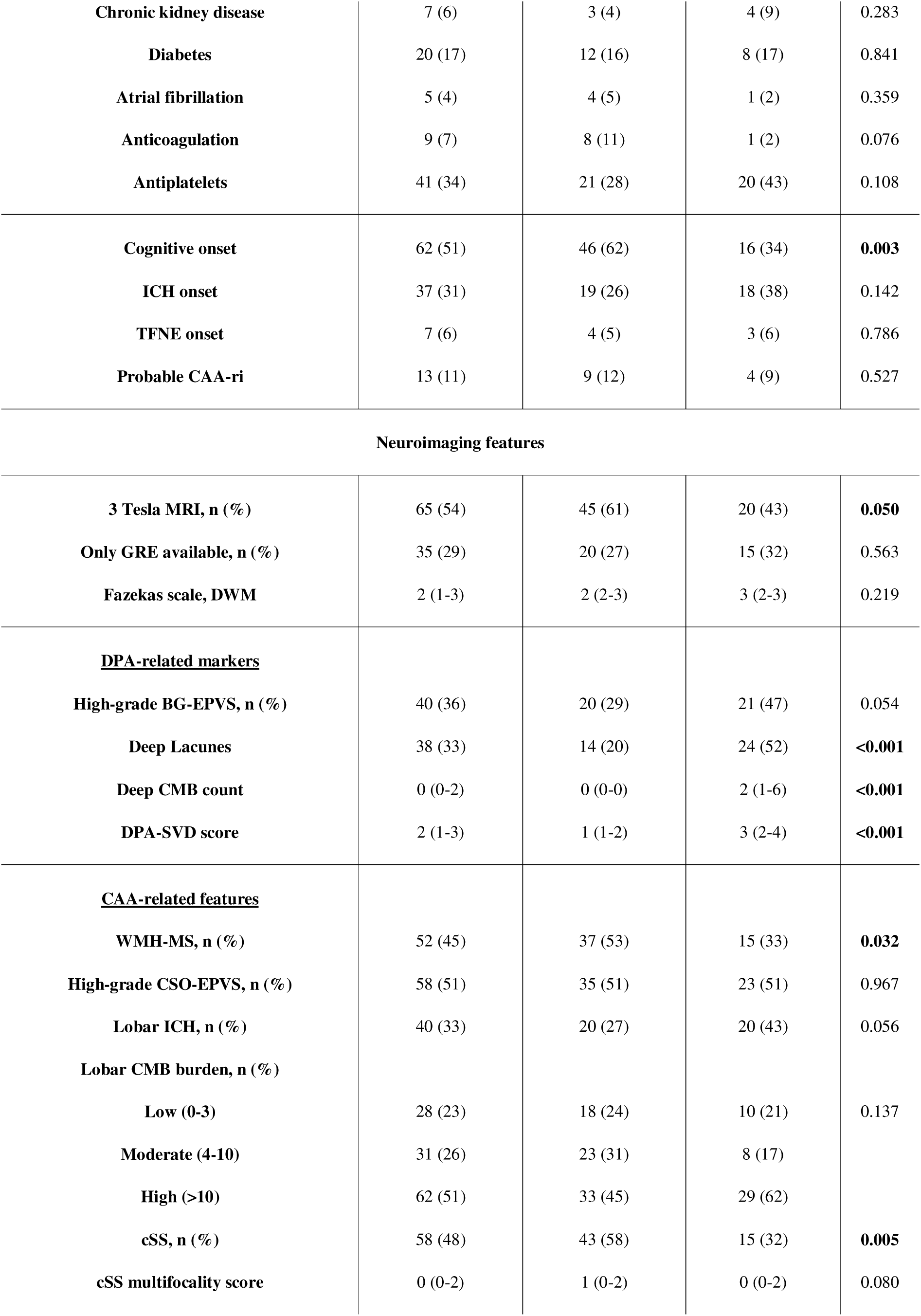
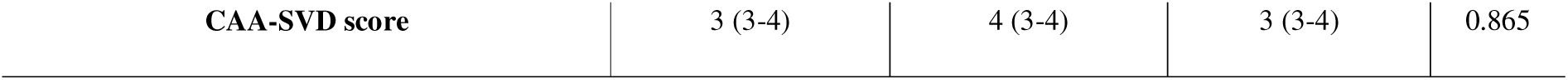
Characteristics of the study participants. Values are expressed as mean±SD or median (IQR), as appropriate, and CSF protein levels in pg/ml. Fazekas scale ranges from 0 to 3. Multifocality scores for cSS range from 0 to 4. Significant p-values are reported in **bold**. Legend: Aβ42=amyloid β 1-42; Aβ40=amyloid β 1-40; AD=Alzheimer’s disease; CAA=cerebral amyloid angiopathy; CMB=Cerebral Microbleeds; cSS=cortical superficial siderosis; DWM=Deep White Matter; y=years; ICH=Intracerebral Hemorrhage; IQR=Interquantile Range; MMSE=Mini-Mental State Examination; MRI=Magnetic Resonance Imaging; p-Tau=phosphorylated Tau. SD=standard deviation; Tau=Tau protein; TFNE=Transient Focal Neurological Episode.

| Variable | Suspected CAA<br>(n=121) | Probable CAA<br>(n=74) | Mixed SVD<br>(n=47) | p<br>value |
| --- | --- | --- | --- | --- |
| Sex, male, n (%) | 42 (35) | 50 (68) | 29 (62) | 0.509 |
| Age at MRI, y | 72 (66-77) | 74 (66-78) | 71 (65-75) | 0.088 |
| MMSE at MRI (n=104) | 26 (23-28) | 25 (23-28) | 27 (23-29) | 0.111 |
| Aβ42 | 447 (323-649) | 371 (315-550) | 542 (356-887) | **0.006** |
| Aβ40 | 7,986 (6,275-10,212) | 7,918 (6,344-10,497) | 7,986 (6,342-9,874) | 0.684 |
| Aβ42/40 ratio | 0.05 (0.04-0.10) | 0.05 (0.04-0.06) | 0.09 (0.05-0.10) | **<0.001** |
| T-tau | 423 (285-665) | 453 (306-673) | 405 (229-650) | 0.476 |
| pTau181 | 48 (29-71) | 60 (38-80) | 32 (22-58) | **<0.001** |
| **Cardiovascular risk factors, n (%)** |  |  |  |  |
| Hypertension | 95 (79) | 54 (73) | 41 (87) | 0.063 |
| Dyslipidemia | 74 (61) | 48 (65) | 26 (55) | 0.293 |
| Smoking | 29 (24) | 16 (21) | 13 (28) | 0.448 |
| **Chronic kidney disease** | 7 (6) | 3 (4) | 4 (9) | 0.283 |
| **Diabetes** | 20 (17) | 12 (16) | 8 (17) | 0.841 |
| **Atrial fibrillation** | 5 (4) | 4 (5) | 1 (2) | 0.359 |
| **Anticoagulation** | 9 (7) | 8 (11) | 1 (2) | 0.076 |
| **Antiplatelets** | 41 (34) | 21 (28) | 20 (43) | 0.108 |
| **Cognitive onset** | 62 (51) | 46 (62) | 16 (34) | **0.003** |
| **ICH onset** | 37 (31) | 19 (26) | 18 (38) | 0.142 |
| **TFNE onset** | 7 (6) | 4 (5) | 3 (6) | 0.786 |
| **Probable CAA-ri** | 13 (11) | 9 (12) | 4 (9) | 0.527 |
| Neuroimaging features |  |  |  |  |
| **3 Tesla MRI, n (%)** | 65 (54) | 45 (61) | 20 (43) | **0.050** |
| **Only GRE available, n (%)** | 35 (29) | 20 (27) | 15 (32) | 0.563 |
| **Fazekas scale, DWM** | 2 (1-3) | 2 (2-3) | 3 (2-3) | 0.219 |
| **<u>DPA-related markers</u>** |  |  |  |  |
| **High-grade BG-EPVS, n (%)** | 40 (36) | 20 (29) | 21 (47) | 0.054 |
| **Deep Lacunes** | 38 (33) | 14 (20) | 24 (52) | **<0.001** |
| **Deep CMB count** | 0 (0-2) | 0 (0-0) | 2 (1-6) | **<0.001** |
| **DPA-SVD score** | 2 (1-3) | 1 (1-2) | 3 (2-4) | **<0.001** |
| **<u>CAA-related features</u>** |  |  |  |  |
| **WMH-MS, n (%)** | 52 (45) | 37 (53) | 15 (33) | **0.032** |
| **High-grade CSO-EPVS, n (%)** | 58 (51) | 35 (51) | 23 (51) | 0.967 |
| **Lobar ICH, n (%)** | 40 (33) | 20 (27) | 20 (43) | 0.056 |
| **Lobar CMB burden, n (%)** |  |  |  |  |
| **Low (0-3)** | 28 (23) | 18 (24) | 10 (21) | 0.137 |
| **Moderate (4-10)** | 31 (26) | 23 (31) | 8 (17) |  |
| **High (>10)** | 62 (51) | 33 (45) | 29 (62) |  |
| **cSS, n (%)** | 58 (48) | 43 (58) | 15 (32) | **0.005** |
| **cSS multifocality score** | 0 (0-2) | 1 (0-2) | 0 (0-2) | 0.080 |
| CAA-SVD score | 3 (3-4) | 4 (3-4) | 3 (3-4) | 0.865 |

### 3.2 Reference CSF biomarker and data-driven cut-off

A bimodal distribution was evident only in the Aβ42/40 ratio, while no clear biological subgroups were visually identified in Aβ42 and Aβ40 alone (**eFigure2**). The associations between z-normalized CSF biomarker levels and MRI-visible manifestations of cSVD are shown in **eTable1**. To summarize, the strongest associations were found between cSVD imaging markers and the Aβ42/40 ratio, showing consistent negative associations with CAA-related features (CAA-SVD score: aOR=0.36 [95%CI 0.24-0.53], p<0.001) and positive associations with DPA-related markers (DPA-SVD score: aOR=3.55 [95%CI 2.40-5.41], p<0.001). These associations were less evident for Aβ42 and significant only with cSS for Aβ40.

We performed a GMM on Aβ42/40 ratio values of the whole cohort to identify a data-driven biological threshold for amyloid positivity (A^+^). The GMM analysis identified an optimal intersection point between these two distributions (0.083 [95%CI: 0.078-0.089] – **eFigure3**). Based on this data-driven cut-off, 32% of the study population (n=39) were classified as A-, while 68% (n=82; 81% of the probable CAA and 45% of the mixed SVD) were classified as A+, with a gain of three patients compared to the AD-related standard cut-off. Applying the same model to Aβ42 alone, we identified a data-driven cut-off (639 [95%CI 516-751] pg/ml; **eFigure3**), with a greater relative uncertainty of the threshold compared to the ratio (13.8% for the Aβ42/40 ratio vs. 36.8% for Aβ42). We observed numerically more patients falling within a diagnostic “gray zone” (95%CI of the data-driven cut-off for amyloid positivity) considering Aβ42 (n=22; 18.2% of the cohort) compared to the Aβ42/40 ratio (n=2; 1.7% of the cohort) as the reference CSF biomarker (**Figure 2**).

**Figure 2.**
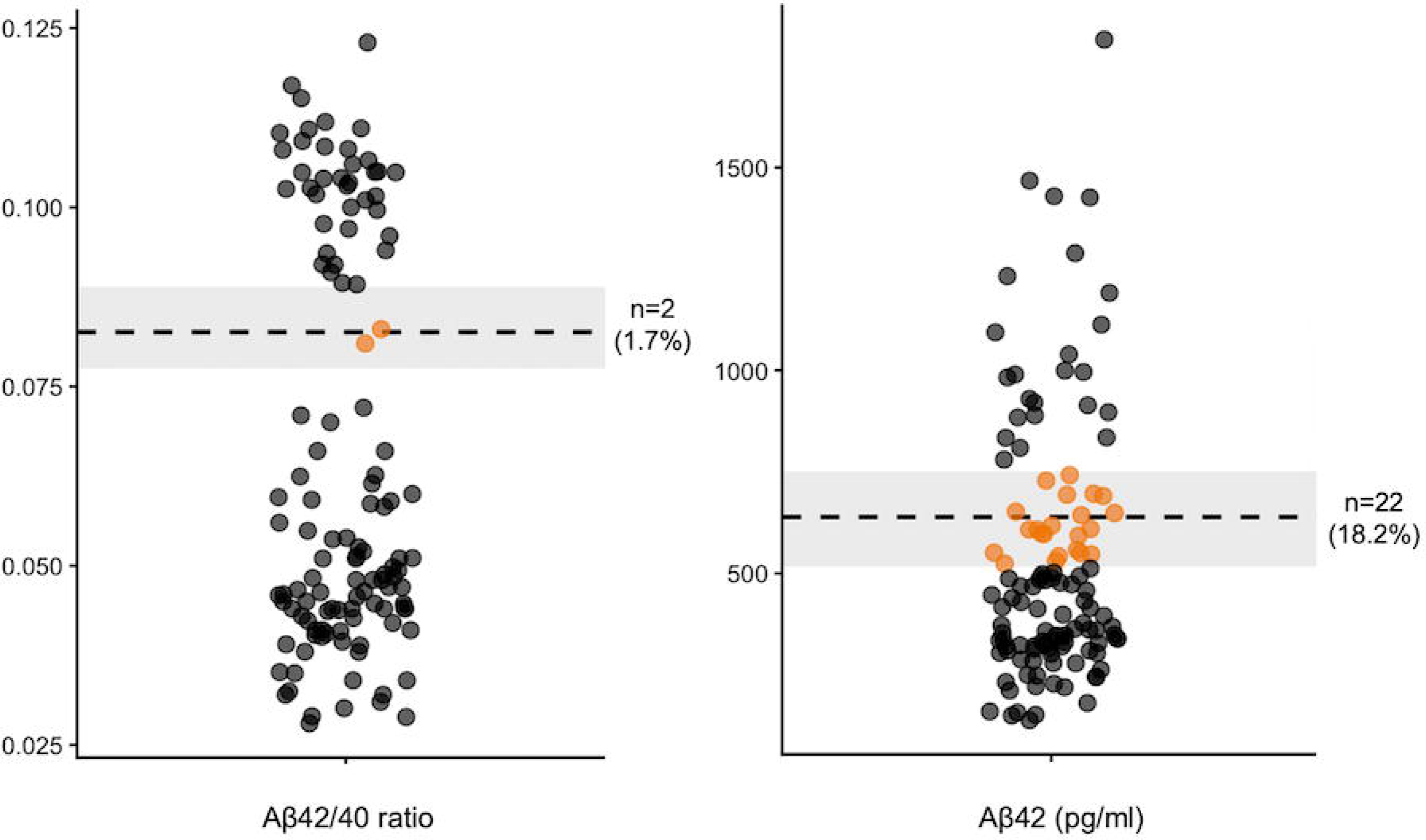
Comparison of the distribution between Aβ42/40 ratio and Aβ42 alone in a suspected CAA cohort and subjects falling in the diagnostic gray zone. Jitter plots illustrating the distribution of patients (n=121) relative to the data-driven cut-off of amyloid-positivity (horizontal dashed lines) and their respective 95% confidence intervals (shaded gray areas). Patients falling within the shaded uncertainty bands are highlighted in orange. (Left) The CSF Aβ42/40 ratio limits diagnostic ambiguity to just 2 patients (1.7%). (Right) Aβ42 leaves 22 patients (18.2%) within this gray zone, demonstrating the possible superior classification performance achieved by the ratio. Legend: Aβ42=amyloid β 1-42; Aβ40=amyloid β 1-40; CAA=cerebral amyloid angiopathy; CSF=Cerebrospinal Fluid;

### 3.3 Incremental Value of CSF Biomarker-Integrated Classification

The integration of amyloid biomarker status substantially modified patient classification. Twenty-two patients (47%) with mixed SVD showed evidence of Aβ deposition and were classified as Aβ-positive mixed SVD, whereas 14 patients (19%) with probable CAA lacked evidence of Aβ deposition and were classified as Aβ-negative probable CAA (**Figure 3)**. Overall, the biomarker-integrated approach successfully reallocated patients, providing a better phenotypical discrimination between the resulting subgroups compared with the current MRI-only classification (**Figure 4 –** for further detail, see **eFigure4)**

**Figure 3.**
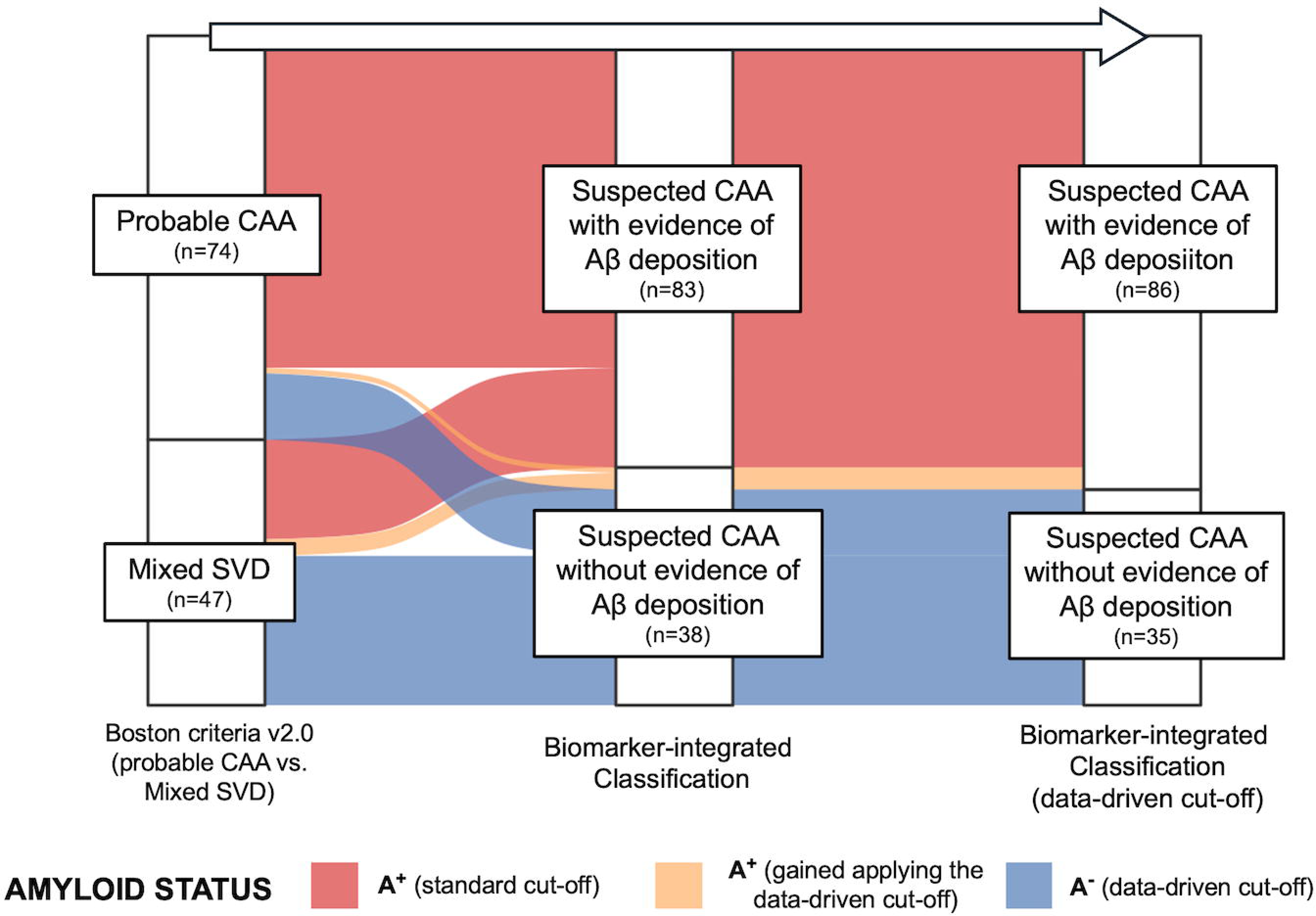
Diagnostic reclassification of patients with suspected Cerebral Amyloid Angiopathy. The alluvial plot illustrates the diagnostic trajectories of the 121 enrolled patients across different classification frameworks. The left axis represents the baseline radiological categorization based solely on the Boston criteria v2.0 (probable CAA vs. mixed SVD). The right axis shows the biomarker-integrated classification applying the data-driven cut-off for amyloid positivity. The central axis depicts the biomarker-integrated classification utilizing the standard Alzheimer’s disease-related cut-off. Red: patients identified as amyloid-positive (A^+^) by the standard threshold; Blue: amyloid-negative (A^-^) individuals according to the data-driven threshold; Orange highlights the subset of patients who gained an A^+^ status through the application of the data-driven cut-off. Legend: CAA=cerebral amyloid angiopathy; SVD=Small Vessel Disease;

**Figure 4.**
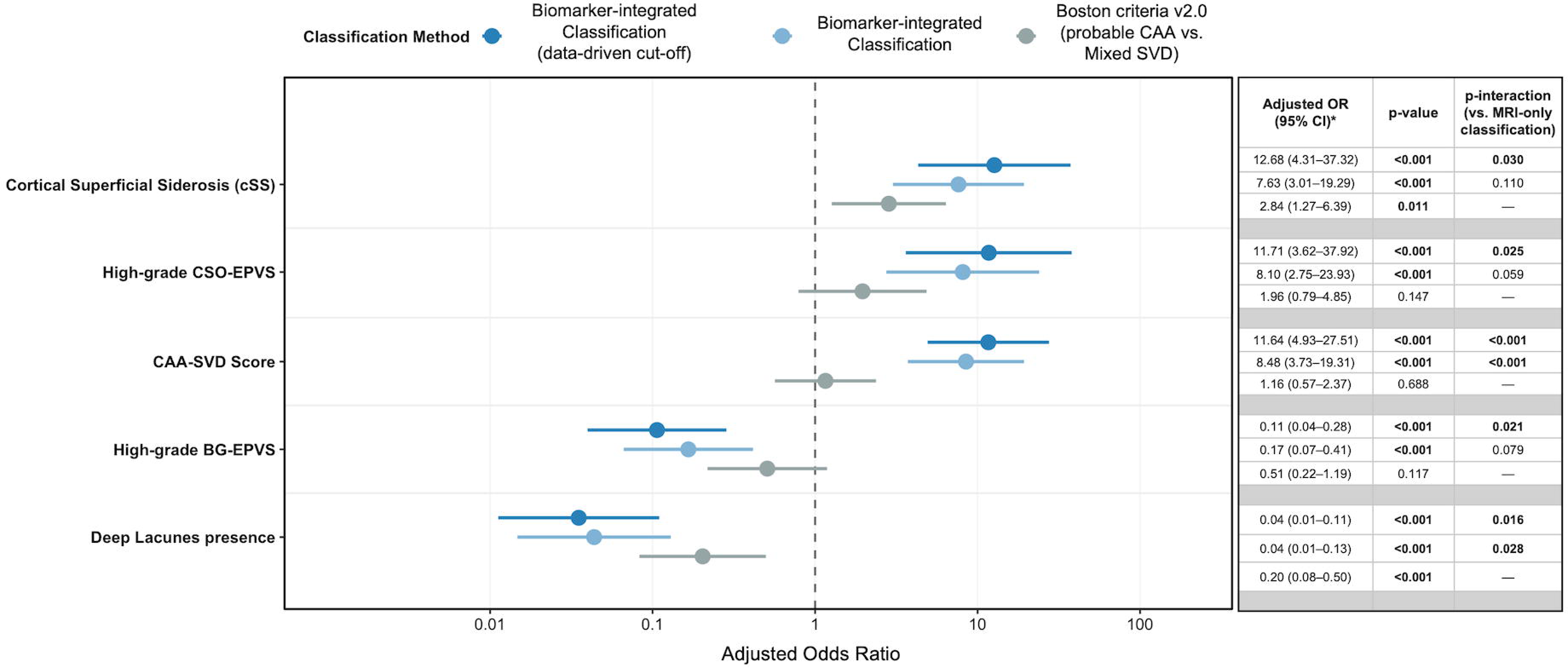
Associations between classifications and MRI-visible manifestation of SVD. The forest plot illustrates the adjusted odds ratios (ORs) and 95% confidence intervals (CIs) for the associations between key radiological markers (cSS, high-grade CSO-EPVS, CAA-SVD score, high-grade BG-EPVS, and deep lacunes) and three distinct classification frameworks. The x-axis is displayed on a logarithmic scale. The right panel reports the exact adjusted ORs, corresponding p-values, and the p-interaction, which formally tests whether the strength of the association is significantly superior when using the biomarker-integrated models compared to the baseline MRI-only classification. Models were ordinal or binomial logistic regression adjusted for relevant confounders [age, center and MRI protocol]. Legend: cSS, cortical superficial siderosis; CSO-EPVS, centrum semiovale enlarged perivascular spaces; CAA, cerebral amyloid angiopathy; SVD, small vessel disease; BG-EPVS, basal ganglia enlarged perivascular spaces; OR, odds ratio; CI, confidence interval.

In detail, MRI-visible CAA markers showed a stronger association with the CSF biomarker-integrated classification (i.e., the Aβ-positive status), particularly when the data-driven cut-off was used instead of the AD-derived cut-off. In particular, the association with cSS was higher in the biomarker-integrated classification (A^+^ vs. A^-^: 64.6% vs. 12.8%, aOR=12.68 [95%CI: 4.31–37.32], p<0.001) compared with the radiological-only framework (probable CAA vs. mixed SVD: 58.1% vs. 31.9%; aOR=2.84 [95%CI: 1.27–6.39], p=0.011 - test for interaction between the two classification methods: p=0.030). The same trend was observed for high-grade CSO-EPVS (probable CAA vs. mixed SVD: 50.7% vs. 51.1%; aOR = 1.96 [95% CI: 0.79–4.85], p=0.147; A^+^ vs. A^-^: 63.2% vs. 26.3%; aOR=11.71 [95%CI: 3.62–37.92], p<0.001 - p for interaction: p=0.025) and the CAA-SVD score (probable CAA vs. mixed SVD: aOR=1.16 [95%CI: 0.57–2.37], p=0.688; A^+^ vs. A^-^: aOR=11.64 [95%CI: 4.93–27.51], p<0.001 – p for interaction: p<0.001). All patients who experienced TFNE as the index diagnostic event were classified as A^+^ (A^+^ vs. A^-^: 9% vs. 0%; p=0.060). To verify their independent associations of radiological features with amyloid positivity, we performed a multivariable logistic regression model including all the known predictors of CAA neuropathology as explanatory variables and the data-driven amyloid status as the dependent variable (**Table 2)**.

**Table 2.** Multivariable logistic regression of MRI markers’ association with evidence of Aβ deposition in the whole cohort. The Aβ42/40 ratio (data-driven cut-off obtained from GMM) was considered the reference for Aβ deposition. Legend: CAA=cerebral amyloid angiopathy; CMB=Cerebral Microbleeds; CSO-PVS=perivascular spaces in the centrum semiovale, cSS=cortical superficial siderosis; OR (95% CI) = odds ratio (95% confidence interval), WMH-MS=white matter hyperintensities in a multispot pattern

| MRI marker | OR (95% CI) | p-value |
| --- | --- | --- |
| $\geq 2$ strictly lobar CMB | 10.4 (3.2-34.4) | **<0.001** |
| $\geq 1$ foci of cSS | 24.1 (5.7-102.3) | **<0.001** |
| Severe CSO-PVS | 5.2 (1.6-16.7) | **0.005** |
| WMH-MS | 0.5 (0.2-1.7) | 0.285 |

By contrast, markers known to be linked to DPA were highly prevalent in the A^-^ subgroup. The association with high-grade BG-EPVS was greater for the biomarker-integrated compared with the standard MRI-based classification (probable CAA vs. mixed SVD: 29.0% vs. 46.7%; aOR=0.51 [95%CI: 0.22–1.19], p=0.117; A^+^ vs. A^-^: 21.1% vs. 65.8%; aOR=0.11 [95%CI: 0.04–0.28], p< 0.001 – p for interaction: p=0.021), as for the deep lacunes (probable CAA vs. mixed SVD: 20.0% vs. 52.2%; aOR=0.20 [95%CI: 0.08–0.50], p<0.001; A^+^ vs. A^-^: 11.7% vs. 74.4%, aOR=0.04, [95%CI: 0.01–0.11], p<0.001 – p for interaction: p=0.016). An inverse relationship was observed between the deep CMB count and the prevalence of amyloid positivity (**Figure 5**). In the subset of patients with suspected CAA and no or low deep CMB burden, the majority were classified as A^+^ (81.3% and 80.0%, respectively). On the contrary, in patients with a moderate burden of deep CMB (n=18), the proportion of A^+^ individuals dropped to 44.4%, and in patients with a severe deep CMB burden (n=13), amyloid positivity became extremely rare (7.7%). Suspected CAA patients classified as A^+^ had never more than 4 deep CMB.

**Figure 5.**
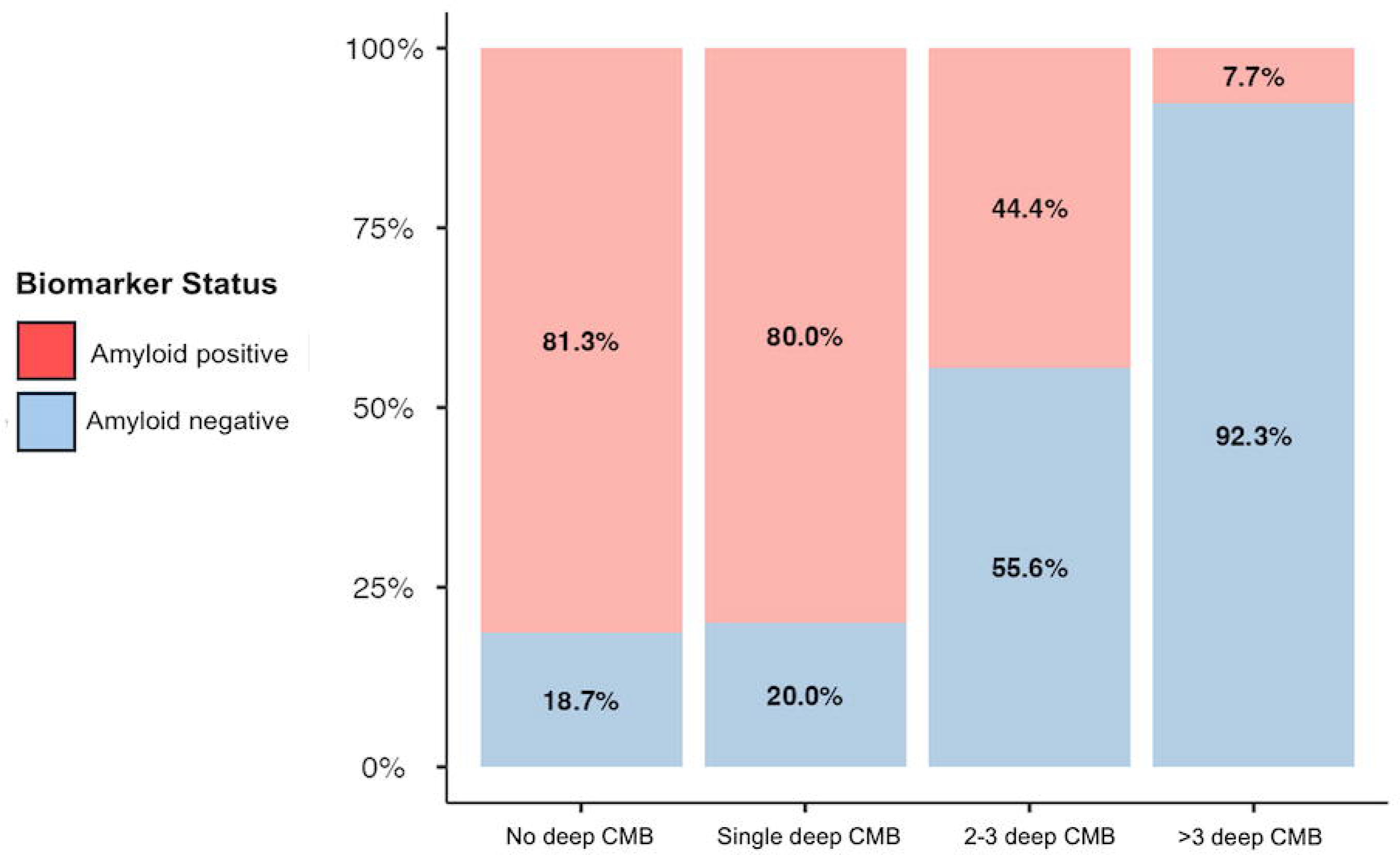
Distribution of amyloid status stratified by deep microbleeds burden. The stacked bar chart illustrates the relative proportion of amyloid-positive (red) and amyloid-negative (blue) patients across categories of deep CMB burden. An increasing number of deep CMBs is associated with a lower prevalence of amyloid positivity. Legend: CMB, cerebral microbleed.

### 3.4 Sensitivity analysis

Results remained consistent after excluding patients with evidence of AD-related tauopathy (i.e., T^+^ patients; n=53), pointing toward the independence of the results from AD co-pathology (**eFigure5**). In a second sensitivity analysis, we excluded patients with CAA-ri (n=13), possible/probable iatrogenic CAA (n=5), and patients with the lumbar puncture performed less than a month from a symptomatic ICH (n=9; final sample size: n=99), obtaining a similar incremental value of the biomarkers-integrated classification (data not shown).

## 4. Discussion

In this multicentre study, we showed that integrating CSF biomarkers (i.e., Aβ deposition evidence) into the classification of patients with suspected CAA may substantially improve the phenotypical concordance with the suspected predominant underlying SVD. These findings were particularly evident when applying the Aβ42/40 ratio, which demonstrated more reliable performance than Aβ42 alone. These data may pave the way for a more standardized application of fluid biomarkers in the classification process of suspected CAA, potentially providing in the future an incremental diagnostic value over the current MRI-based classification.

CSF biomarkers provide direct biological evidence of Aβ deposition and are recommended as a first-line test to support the diagnosis of AD in patients with a consistent clinical picture^30^. In this context, the Aβ42/40 ratio is regarded as the standard for determining the amyloid (“A”) status over Aβ42, given its superior correspondence with amyloid PET and its capacity to overcome inter-individual variability in amyloid production^31–33^. The observed bimodal distribution of the Aβ42/40 ratio in a cohort of suspected CAA suggests a more reliable distinction in subgroups using the ratio compared to the single Aβ peptides, even in this context. Consequently, the ratio led to a substantial decrease in the occurrence of borderline or “grey zone” values, while Aβ42 suffered from this limit. Furthermore, Aβ42 exhibited a weaker association with the radiological signature of CAA than the Aβ42/40 ratio, suggesting a potential enhanced efficacy related to the normalization process on Aβ40 levels. On the other hand, Aβ40 levels did not produce similar results, which is unexpected given its established role as the predominant Aβ species with vascular deposition^12,34^. Notably, the absence of difference in Aβ40 levels comparing probable CAA and mixed SVD was previously reported, and the selection of suspected CAA with mixed SVD could have reduced a possible difference between the two groups^35^. Nevertheless, a role for Aβ40 levels seems to be potentially present, representing probably a marker of pro-hemorrhagic CAA rather than a marker of amyloid status, given its association with cSS and hemorrhagic risk^15,36–39^. This is of particular clinical interest, given the paucity of robust prognostic indicators in CAA, with the exception of cSS^40^.

The role of CSF biomarkers in the CAA field is progressively emerging, and the current pathophysiological framework supports it as the earliest demonstrable biological process in vivo in the genetic form of the disease^41,42^. If we consider that a low level of Aβ species in the CSF can effectively identify CAA, the application of a cut-off derived from the AD field may be considered arbitrary. The observation that our data-driven cut-off, derived in an unsupervised way, closely approximates established AD-related thresholds provides indirect validation for our findings; however, it is important to note that an optimization process should be addressed in future studies with a proper neuropathological reference. Despite this, the subtle yet discernible discrepancy between the two cut-offs, likely attributable to a relative reduction of Aβ40 in CAA compared to AD, holds significant clinical relevance at the individual subject level, given the gain of three patients (3.5% of the amyloid-positive population) classified as A^+^ compared to the application of the standard cut-off^15^.

The primary finding of this study is the radical divergence of MRI-visible manifestations of SVD following biomarker-integrated reclassification compared to the standard distinction between probable CAA and mixed SVD. Recently, a similar approach was applied using amyloid PET, which demonstrated a role in aiding the identification of patients with high hemorrhagic risk^10^. Selecting for this study patients with “suspected CAA” (”probable CAA” according to the Boston criteria v2.0, but without considering deep bleeding as an exclusion criterion), we positioned ourselves within the differential diagnosis of the two main age-related SVD (i.e., CAA and arteriosclerosis)^1,3^. The MRI-only classification demonstrated significant overlap between the baseline radiological phenotypes; this is likely attributable to the inclusion criteria of the study, which did not encompass the full range of mixed SVD. The application of a biological classifier successfully reallocated ambiguous patients, as evidenced by the purification of the radiological phenotype between biological subgroups, with CAA hallmarks (i.e., cSS, CSO-EPVS, CAA-SVD score) strongly anchored to amyloid positivity and DPA-related markers with amyloid negativity.

A significant confounding factor in research on CAA biomarkers is the frequent pathological concurrence with AD, which diminishes confidence in the pathology being measured in individual patients with CSF (or plasma) biomarkers^43,44^. With the exclusion of patients with evidence of Alzheimer’s-related tauopathy (T+) in the sensitivity analysis, we confirmed that our findings are relatively independent of AD copathology. This finding indicates that the evidence of CSF reduction in Aβ peptides is not merely an epiphenomenon of concomitant AD, but rather independently reflects the process of vascular amyloid deposition. However, even if this is true at the group level, the clinical context should always guide the interpretation of the biomarkers at the single-subject level, given the substantial overlap between age-related pathologies^45^.

Deep CMB and the DPA-SVD score were not considered in the comparison between classifications to avoid circularity with the definition of mixed SVD. However, the analysis of the deep CMB count in relation to amyloid status provided relevant information. The prevalence of evidence of Aβ deposition was 45% in patients with mixed SVD, a significantly lower percentage than in patients with probable CAA (82%; p<0.001), a difference that supports the approach of the Boston Criteria v2.0, which aims to preserve a high diagnostic specificity^3^. Our observation that a high burden of deep CMBs (over 3) was very unlikely associated with an amyloid-positive status (7.7%), while patients with low or moderate burden had a greater prevalence of amyloid-positive patients (80% and 44%, respectively), suggests that the pathological driver of mixed SVD is heavily variable. Although arteriosclerosis is likely the predominant underlying cSVD in most mixed SVD cases, the co-presence of CAA should be considered in patients with a relatively low hemorrhagic deep burden^7,8^. The confirmation of a biological process consistent with Aβ deposition would potentially represent an indicator of concomitant CAA in this context, along with the presence of cSS^46^. This could represent in the future a formal clinical indication for CSF examination, in order to clarify the predominant SVD and the subsequent prognosis.

### 4.1. Limitations of the study

This study has some limitations. First, the absence of the diagnostic gold standard for CAA (i.e., neuropathology) excluded the possibility of directly verifying the diagnostic accuracy of CSF biomarkers in this context. Nevertheless, the trend of the radiological divergence from the current classification to the data-driven biomarker-integrated one is suggestive of better phenotypical discrimination. Second, the sample size is relatively small, with low prevalence in the cohort of certain subgroups of patients (e.g., patients with TFNE, mixed SVD with high burden of deep CMB). Third, the retrospective nature of the data collection may introduce bias. Although CSF analysis was systematically proposed during the study period, selection bias was intrinsic in specific clinical situations (e.g., patients with severe ICH and poor short-term prognosis, who rarely undergo MRI scanning). Fourth, despite the statistical adjustments for scanner field strength and sequence type, clinical MRIs are inherently characterized by heterogeneous neuroimaging protocols. However, all key sequences necessary to assess radiological features were included. The CMB count is highly dependent on the MRI protocol; thus, the threshold of deep CMB above which we did not observe amyloid-positive patients should be considered an exploratory result, and future studies should compare the performance of other metrics (such as lobar-to-deep ratio of CMB). Finally, even if models were corrected for center, all these findings are currently cohort-specific and should be validated in external cohorts.

## 5. Conclusion

In conclusion, the findings of this proof-of-concept study suggest that the evidence of Aβ deposition may offer valuable insights for the classification of patients with suspected CAA. This biomarker-integrated approach has the potential to reduce diagnostic ambiguity and to establish the presence (rule in) or absence (rule out) of CAA in appropriate contexts, such as in challenging cases and mixed SVD. In a manner analogous to the ATN framework within the AD field, the integration of an in vivo neuropathological proxy of CAA may provide a standardised and objective supportive biomarker in clinical practice. Further research is required to validate the incremental value of an integrated biological-radiological framework to enhance diagnostic accuracy in CAA.

## Acknowledgment

AU an FM were supported by #NEXTGENERATIONEU (NGEU) and funded by the Ministry of University and Research (MUR), National Recovery and Resilience Plan (NRRP), project MNESYS (PE0000006) – (DN. 1553 11.10.2022). F.P was supported by PRIMARIA inside the MNESYS framework CUP: B33C22001060002 – NextGenerationEU - M4C2)

AS was supported by #NEXTGENERATIONEU (NGEU) and funded by the Ministry of University and Research (MUR), National Recovery and Resilience Plan (NRRP), project RAISE (ECS00000035) – (DN. 1053 del 23.06.2022).

## Author Contributions

ML: study concept and design, data collection, imaging analysis, statistical analysis, write-up. MCR: study concept and design, data collection, imaging analysis, write-up, critical revisions. IG: data collection, critical revisions; FM (Mazzacane): drafting manuscript, critical revisions; BO: drafting manuscript, critical revisions; LL: drafting manuscript, critical revisions; AD: data collection, critical revisions; FM (Massa): data collection, critical revisions; ES: drafting manuscript, critical revisions; LG: imaging analysis, critical revisions. GP: drafting manuscript, critical revisions; VDF: drafting manuscript, critical revisions; AC: drafting manuscript, critical revisions; FB: drafting manuscript, critical revisions; SMG: drafting manuscript, critical revisions; MGK: drafting manuscript, critical revisions; FP: drafting manuscript, critical revisions; AU: drafting manuscript, critical revisions; AS: drafting manuscript, critical revisions; MDS: drafting manuscript, critical revisions; LMF: imaging analysis, critical revisions; LR: study concept and design, imaging analysis, critical revisions; MP (Pardini): study concept and design, writeup, critical revisions. All authors have read and agreed to the published version of the manuscript.

## Data Availability

The anonymized data and code that support the findings of this study will be made available from the corresponding author upon reasonable request.

## ABBREVIATIONS

AD: Alzheimer’s Disease
AT(N): Amyloid Tau Neurodegeneration Research Framework
BG: Basal Ganglia
CAA: Cerebral Amyloid Angiopathy
CMB: Cerebral Microbleed
CSF: Cerebrospinal Fluid
CSO: Centrum Semiovale
cSS: Cortical Superficial Siderosis
EPVS: Enlarged Perivascular Spaces
GRE: GRadient Echo sequences
ICH: Intracerebral Hemorrhage
MMSE: Mini-Mental State Examination
pTau: phosphorylated Tau
SVD: Small Vessel Disease
SWI: Susceptibility Weighted Imaging
TFNE: Transient Focal Neurological Episode

