## Supplemental materials for "Incremental Value of Cerebrospinal Fluid Biomarker-Integrated Classification of Cerebral Amyloid Angiopathy"

**eFigure 1 - Flowchart of patient selection.**

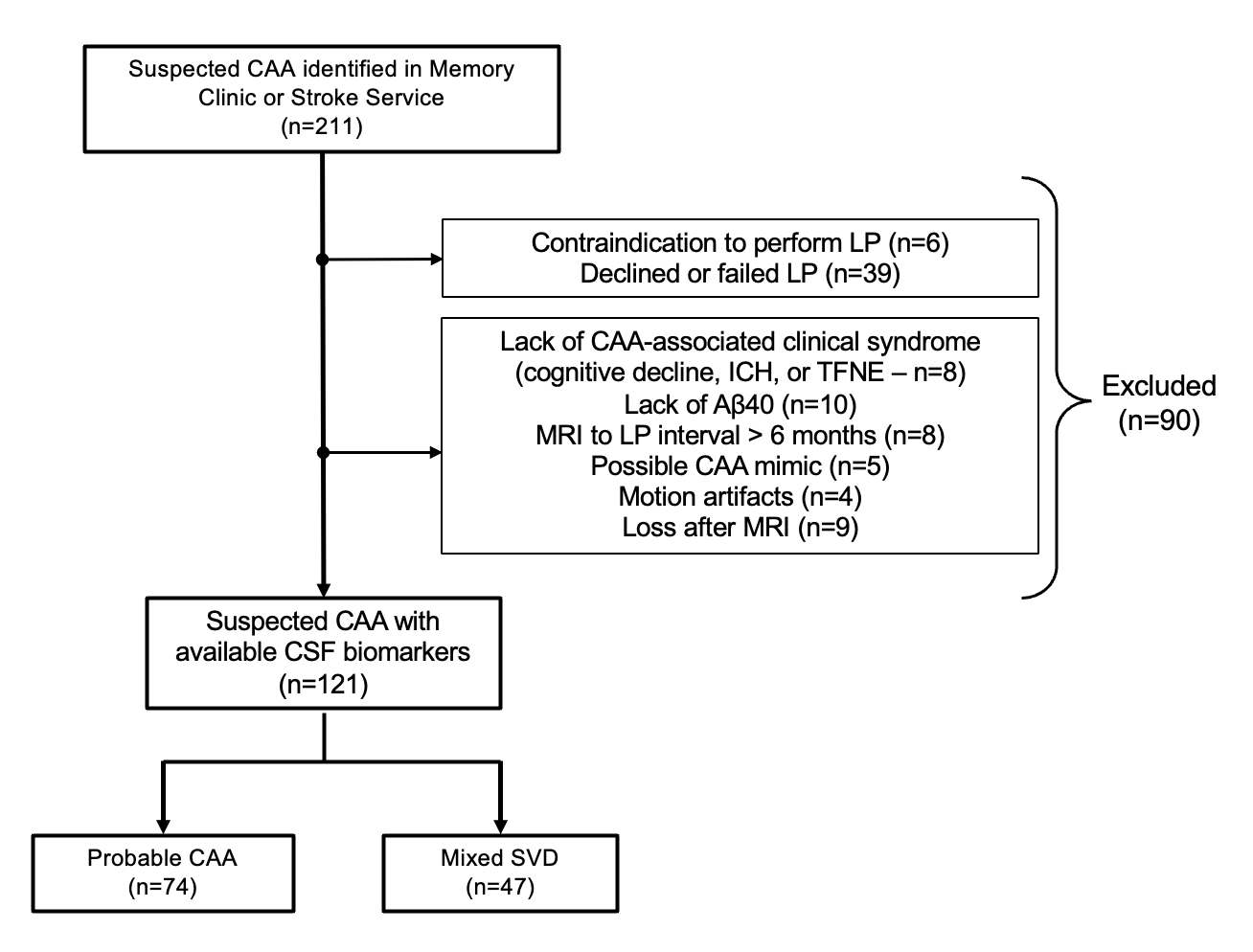
Legend: CAA indicates cerebral amyloid angiopathy; CSF, Cerebrospinal Fluid; ICH, intracerebral hemorrhage; LP, lumbar puncture; MRI, magnetic resonance imaging; SVD, small vessel disease; and TFNE, transient focal neurological episode.

**eFigure 2 - Distribution of CSF biomarker levels.** In the upper row, the density of Aβ42 (on the left), Aβ40 (on the right), and the Aβ42/40 ratio (in the middle) of the whole “suspected CAA” cohort. In the lower row, distribution in the radiological subgroups (probable CAA and mixed SVD). Dragged lines represent the standard AD-related cut-off applied at our centers.

Legend: Aβ42=amyloid β 1-42; Aβ40=amyloid β 1-40; CAA, cerebral amyloid angiopathy; CSF, Cerebrospinal Fluid; SVD, Small Vessel Disease

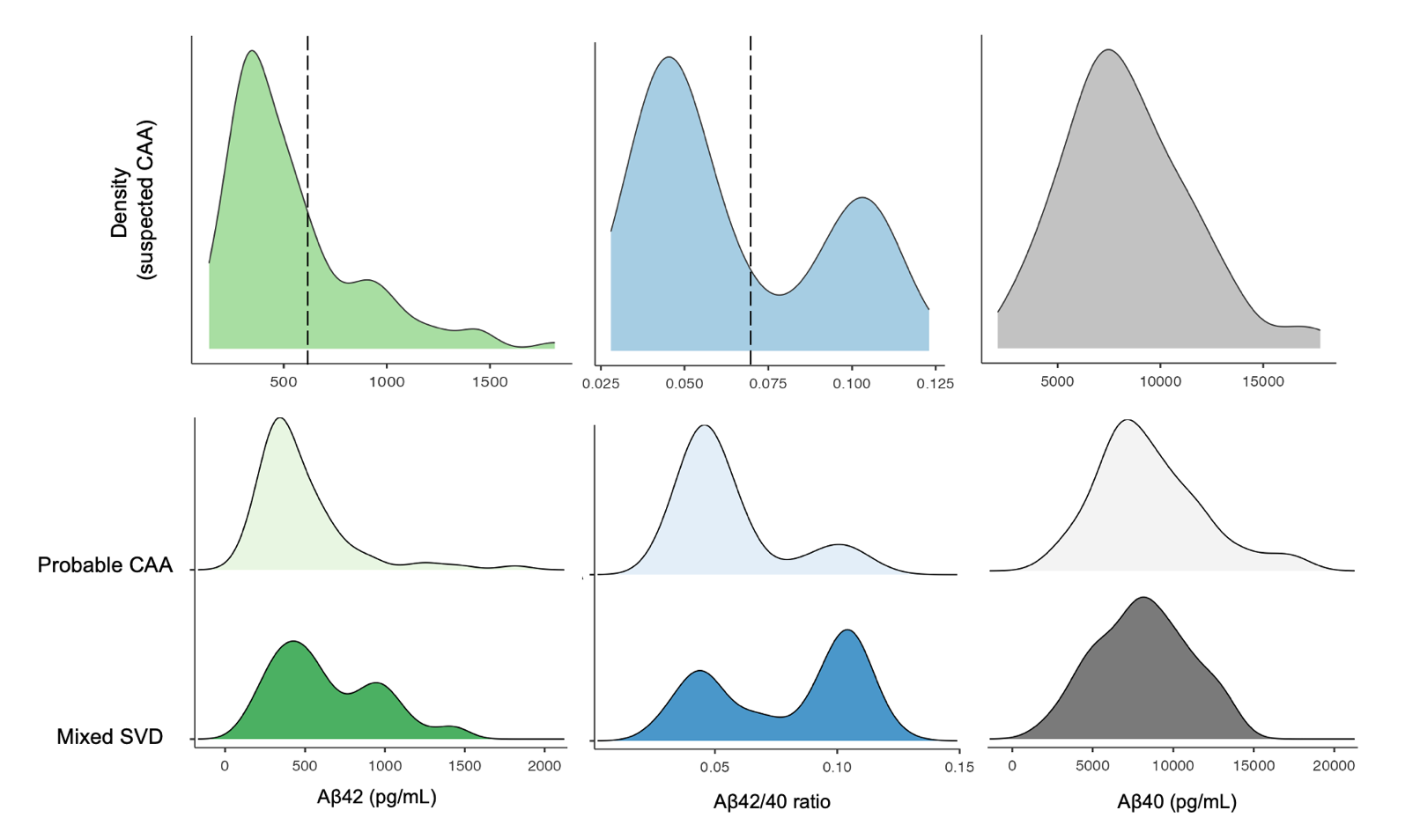

**eFigure 3 - Data-driven identification of Aβ42/40 ratio and Aβ42 cut-off via Gaussian Mixture Modeling (GMM).** GMM applied to the CSF Aβ42/40 ratio and Aβ42 in patients with suspected CAA (n=121). The gray line represents the overall density of the distribution. The red curve illustrates the modeled distribution of the amyloid-positive population (A^+^), while the blue curve represents the amyloid-negative population (A-). The vertical dashed line indicates the data-driven intersection point (light blue area: 95% confidence interval), which is different from the cut-off currently applied for the AD diagnosis at our centers.

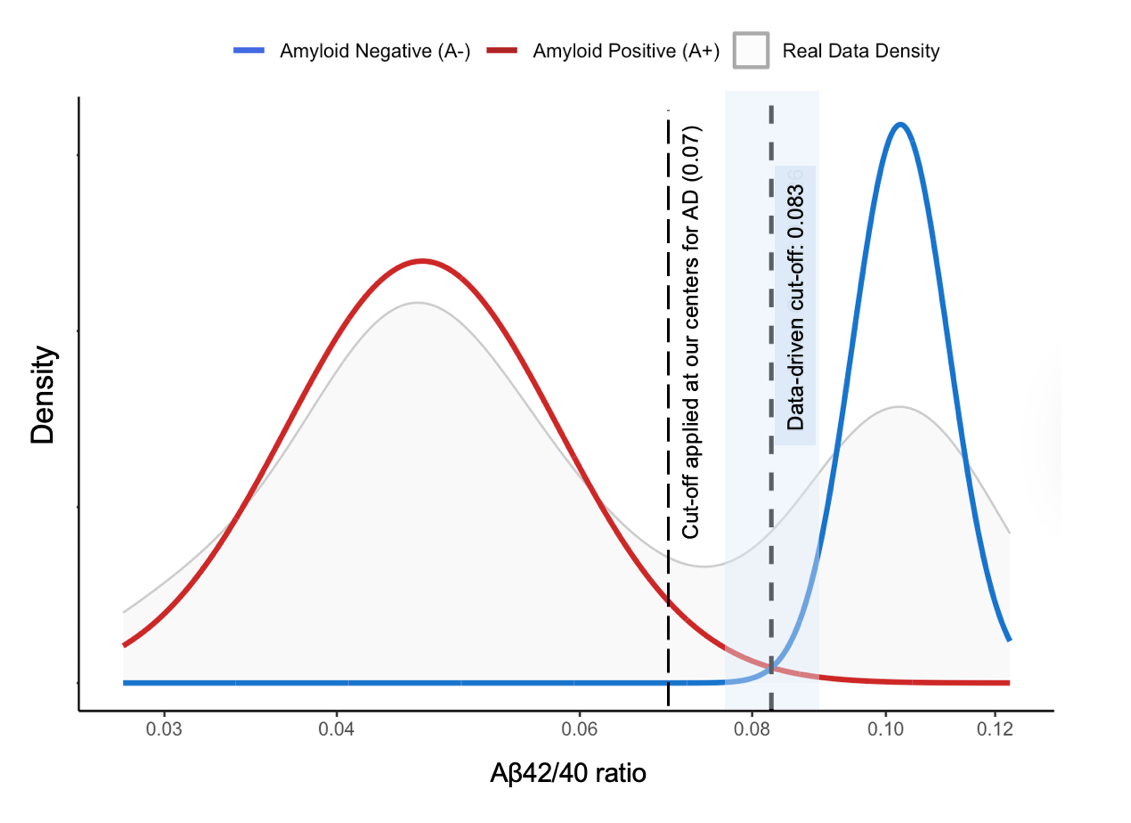

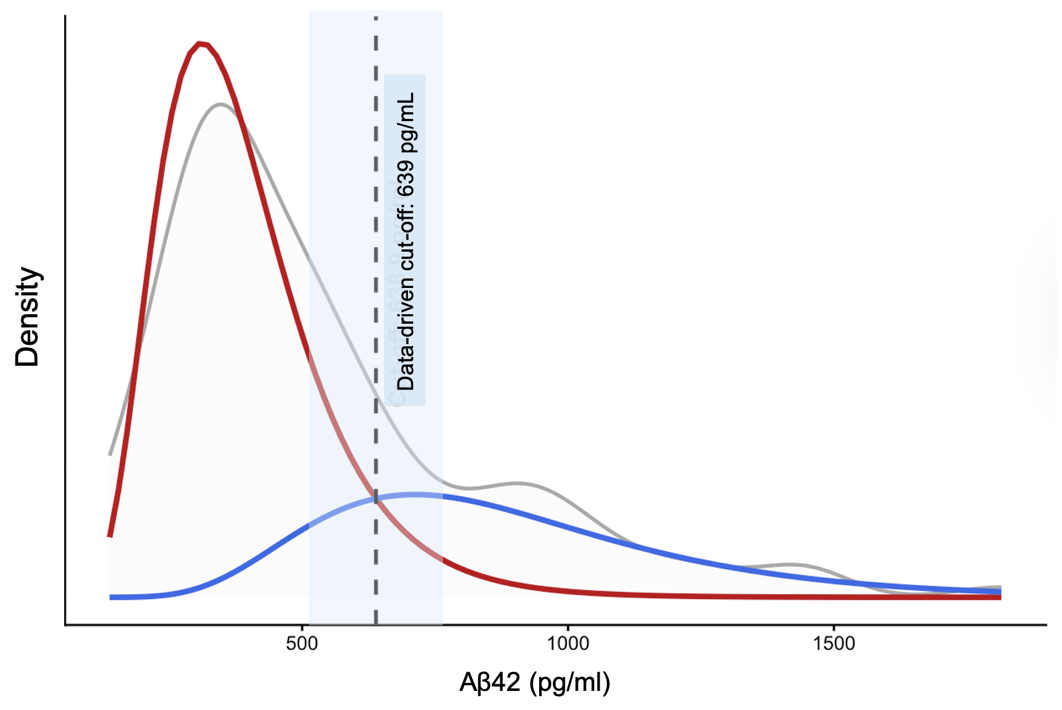
Legend: Aβ42=amylo

**eFigure 4 - Prevalence of CAA- and DPA-related radiological features in MRI-only vs. biomarker-integrated classification of CAA.** OR obtained from binomial or ordinal logistic regression models (adjusted for age, center, and MRI protocol). Legend: Aβ42=amyloid β 1-42; Aβ40=amyloid β 1-40; BG-EPVS: enlarged perivascular spaces of the basal ganglia; CAA=cerebral amyloid angiopathy; CSO-EPVS: enlarged perivascular space of the centrum semiovale; cSS=cortical superficial siderosis; SVD=small vessel disease.

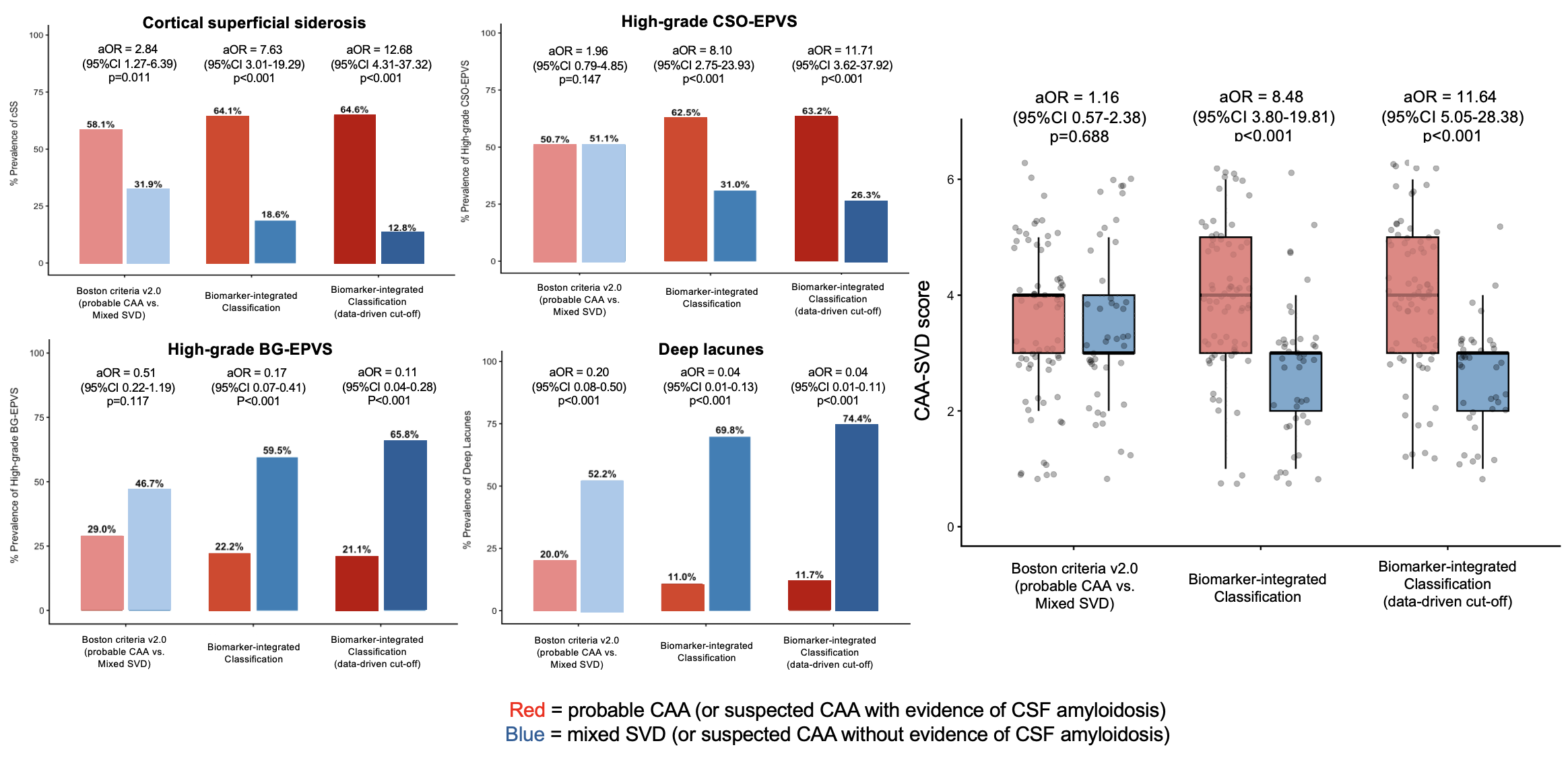

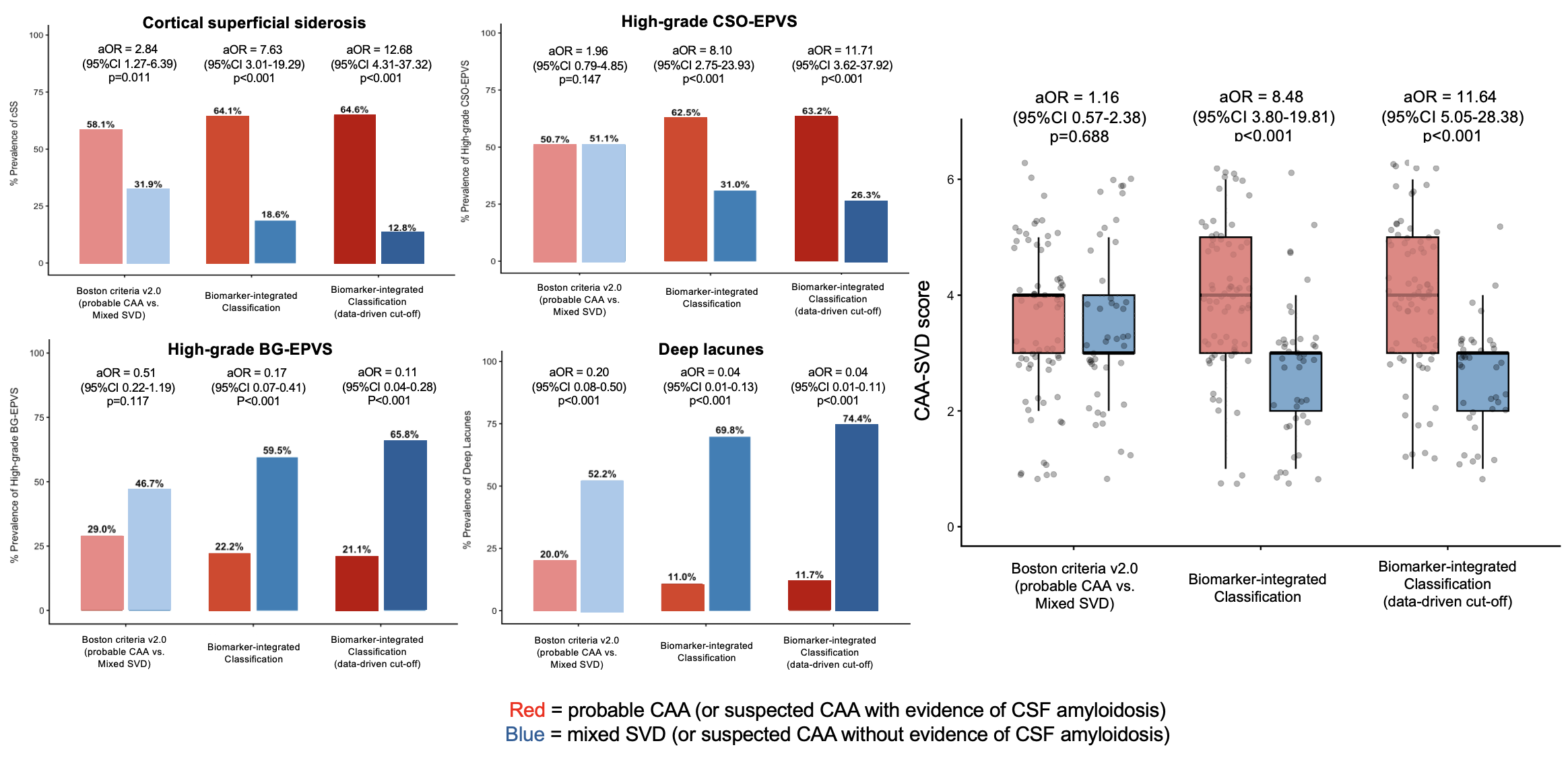

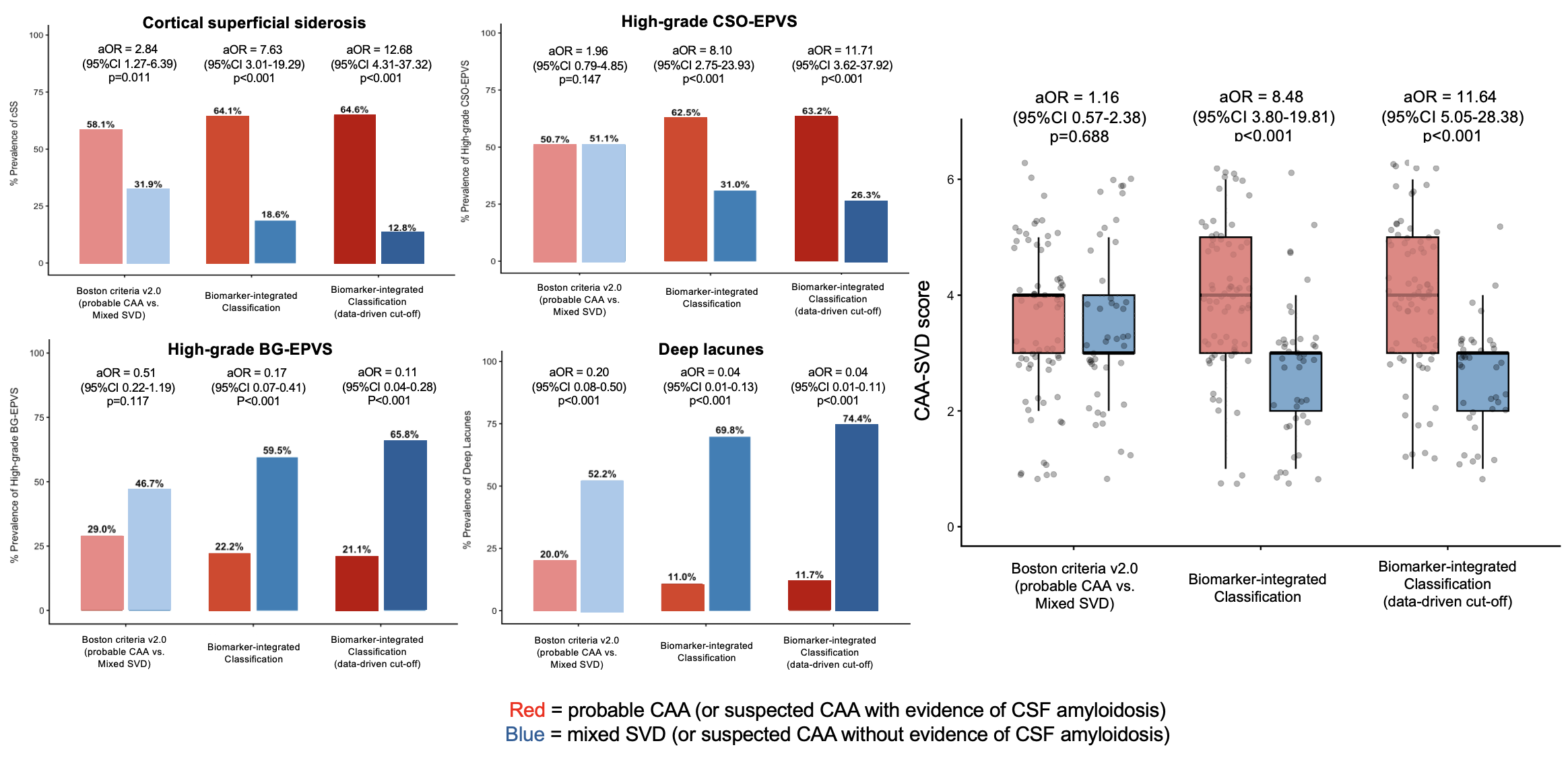

**eFigure 5 - Associations between classifications and MRI-visible manifestation of SVD after the exclusion of patients with evidence of AD-related tauopathy (T+).** The forest plot illustrates the adjusted odds ratios (ORs) and 95% confidence intervals (CIs) for the associations between key radiological markers (cSS, high-grade CSO-EPVS, CAA-SVD score, high-grade BG-EPVS, and deep lacunes) and three distinct classification frameworks. The x-axis is displayed on a logarithmic scale. The right panel reports the exact adjusted ORs, corresponding p-values, and the p-interaction, which formally tests whether the strength of the association is significantly superior when using the biomarker-integrated models compared to the baseline MRI-only classification. Models were ordinal or binomial logistic regression adjusted for relevant confounders [age, center and MRI protocol].

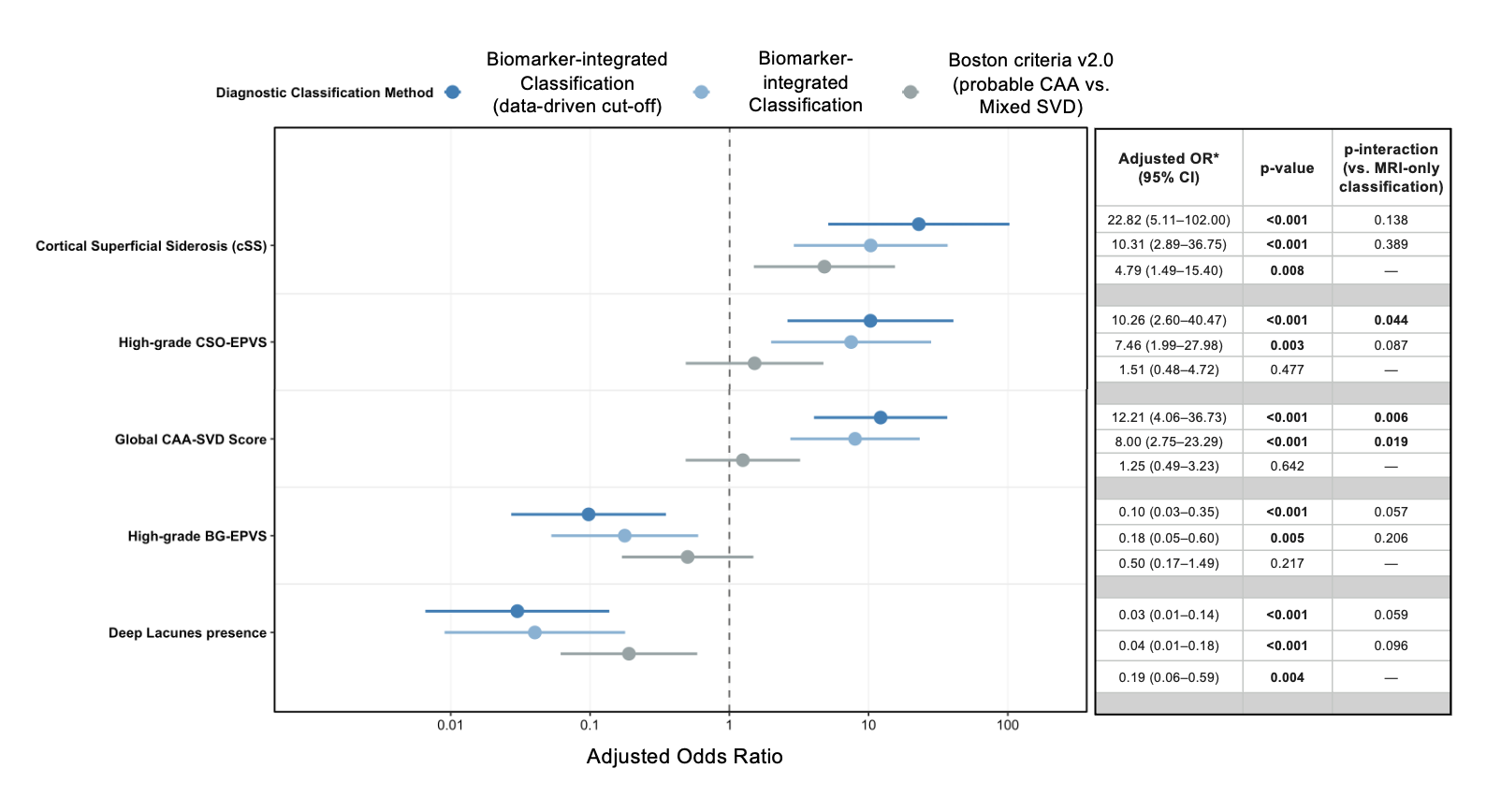
Legend: cSS, cortical superficial siderosis; CSO-EPVS, centrum semiovale enlarged perivascular spaces; CAA, cerebral amyloid angiopathy; SVD, small vessel disease; BG-EPVS, basal ganglia enlarged perivascular spaces; OR, odds ratio; CI, confidence interval.

**eTable 1 - Associations between CSF biomarker levels and MRI-visible manifestations of cSVD.** Effect sizes are expressed as OR (95%CI), obtained from binomial or ordinal logistic regression models, after Z-score normalization of CSF biomarker to obtain comparable effect sizes (models adjusted for age, center, MRI field [1.5 vs. 3 Tesla], and sequence [GRE vs. SWI]; this latter in models considering hemorrhagic features). Color code: yellow cells: p<0.05; orange cells: p<0.01; red cells: p<0.001;

Legend: Aβ42=amyloid β 1-42; Aβ40=amyloid β 1-40; CAA=cerebral amyloid angiopathy; CMB=Cerebral Microbleeds; cSS=cortical superficial siderosis; DPA=Deep Perforating Arteriopathy; DWM=Deep White Matter; ICH=Intracerebral Hemorrhage; MRI=Magnetic Resonance Imaging; p-Tau=phosphorylated Tau.

|  | Neuroimaging markers | Aβ42 | Aβ40 | Aβ42/40  ratio |
| --- | --- | --- | --- | --- |
|  | Fazekas scale  (DWM) | 1.29  (0.91-1.86) | 1.03  (0.73-1.46) | 1.40  (0.98-2.01) |
| **DPA-related features** | EPVS-BG | 2.48  (1.69-3.74) | 1.32  (0.93-1.90) | 2.66  (1.78-4.05) |
|  | Deep lacune  (presence) | 3.38  (1.99-5.75) | 1.08  (0.73-1.62) | 4.91  (2.78-8.66) |
|  | Deep CMB  (count) | 1.71  (1.19-2.52) | 0.94  (0.66-1.36) | 2.70  (1.79-4.16) |
|  | Deep CMB  (presence) | 1.53  (1.03-2.28) | 0.84  (0.57-1.24) | 2.35  (1.51-3.64) |
|  | DPA-SVD score | 2.50  (1.76-3.62) | 1.16  (0.85-1.60) | 3.55  (2.40-5.41) |
| **CAA-related features** | WMH-MS  (presence) | 0.68  (0.46-1.01) | 0.90  (0.61-1.33) | 0.64  (0.43-0.95) |
|  | EPVS-CSO | 0.39  (0.25-0.57) | 0.90  (0.62-1.30) | 0.28  (0.18-0.43) |
|  | cSS  (presence) | 0.25  (0.14-0.44) | 0.49  (0.32-0.76) | 0.38  (0.24-0.60) |
|  | cSS  (multifocality score) | 0.29  (0.18-0.45) | 0.51  (0.36-0.72) | 0.43  (0.28-0.64) |
|  | >2 strictly lobar CMB  (presence) | 0.71  (0.48-1.05) | 1.26  (0.85-1.85) | 0.46  (0.30-0.71) |
|  | CAA-SVD score | 0.39  (0.27-0.56) | 0.71  (0.51-1.01) | 0.36  (0.24-0.53) |
|  | Lobar ICH  (presence) | 0.80  (0.54-1.20) | 0.70  (0.47-1.04) | 1.01  (0.67-1.52) |
